# Assessing the frequency, quantity, and heavy use patterns of marijuana flower among adults with HIV in Florida

**DOI:** 10.1101/2025.03.14.25323993

**Authors:** Donald D Porchia, Yancheng Li, Gladys Ibañez, Gabriela Plazarte, Verlin Joseph, Eric C Porges, Yan Wang, Samuel Wu, Robert L Cook

## Abstract

**Background and Aims:** Measuring the quantity of marijuana flower use is challenging and there is no standardized method of measurement, yet it is critical for cannabis researchers investigating its effects on health outcomes. We sought to identify the frequency and quantity of marijuana flower used per day, the average size of a joint, blunt or bowl, and the average amount of marijuana flower consumed per hit. We also sought to examine the distribution of heavy daily use in terms of grams of flower per day.

**Methods:** As part of the Marijuana Associated Planning and Long-term Effects (MAPLE) longitudinal cohort study, an underrepresented, population of persons with HIV (PWH), who were marijuana flower users (n = 253) (60.1% Age ≥50, 54.4% Male, and 66.4% Black) completed a retrospective, calendar-based timeline follow-back (TLFB) measure. Participants reported on their frequency and quantity of marijuana flower consumed in grams, the number of hits per dose, and methods of consumption during the 30 days prior to each study visit.

**Results:** Of the 253 study participants, 208 (82%) used marijuana flower exclusively and 52% used daily, with a median quantity of 0.8 grams/day. The most common methods of flower consumption and their median quantity consumed were blunts (33%, 1.0 grams), joints (32%, 0.5 grams), and bowls (12%, 0.3 grams). The median amount of marijuana used per hit was 0.063 grams. The proportion who had at least one heavy use day in a month, or heavy use every day of the month was 30% and 6% when heavy use was defined as 3 grams/day, 43% and 13% for 2 grams/day, and 59% and 23% for 1 gram/day.

**Conclusions:** Our results in this underrepresented population of PWH who were marijuana flower users are similar to others in defining the median quantity of a hit, joint and blunt in healthy, young white, male populations. However, the median size of a bowl was smaller than commonly reported. Over half of the sample population consumed greater than 1 gram/day in the previous month and almost a quarter used at least 1 gram of every day of the month.

## 1. Introduction

Cannabis use is widespread and reported to be increasing^1,2^, with 18% of adults in America reporting use in the past year^3^. With such widespread use, it is important to understand the impact of cannabis components such as delta9-THC on health. Marijuana flower, the most commonly used marijuana product, obtained either within or without of the formal medical marijuana system, is commonly used to help manage a variety of symptoms such as chronic pain or nausea and various mental health conditions such as depression, anxiety and post-traumatic stress disorder, as well as recreationally. To better understand the consequences of marijuana use, particularly in the context of dose response relationships, accurate and low burden cannabis consumption metrics are vital for researchers investigating such health outcomes.

The majority of cannabis use is combusted marijuana flower^4,5^ and it is challenging to estimate the quantity consumed. Individual self-reports are both easily measurable and cost effective, but they often provide a less accurate measure of cannabis use^6–11^. Surveys using a single frequency of use measure are still common^12,13^. Yet such a one dimensional measure is not sufficient when trying to estimate the quantity of flower consumed and its impact on health outcomes^14^. Many multi-dimensional surveys have used a TLFB instrument to characterize both the frequency in times per day, and quantity of flower in grams, joints, hits, or even hours of intoxication per day^15–21^. Recent surveys using a unit-preference design have attempted to allow participants to choose the unit in which they wish to report their use such as grams, hits, or joints^22–27^.

However, all such designs are limited by the lack of a standard unit of cannabis use^28,29^. In the literature, it is common to see the size of a “standard” joint, blunt, or bowl being reported as 0.25 or 0.5 grams, 0.5, 1.0 grams or more, and 0.5 grams respectively^18,22,30^. In the case of a hit, the range commonly found in the literature was from 0.01 to .1 gram per hit^22,31–33^. However, these estimates were mainly derived from study populations consisting of young, healthy, white males. Estimates seem lacking for older, underrepresented, populations living with a chronic disease such as HIV. Therefore, our main aim was to try to use a multi-dimensional TLFB survey in order to answer the question: What is the typical size of a joint, blunt, bowl or hit?, in regards to this underrepresented population.

To answer these questions, as part of this longitudinal cohort study of marijuana use in persons with HIV, we sought to use data gathered from a timeline follow back questionnaire to quantify the average size of a joint, blunt or bowl, the average amount of flower used in grams/day and the average amount of marijuana flower per hit. As a secondary aim, we also examined the distribution of heavy use in terms of quantity of flower consumed. Since there is no consensus on the definition of heavy use based on grams of flowers used daily, we chose to examine usage levels at 1, 2 or 3 grams per day.

## 2. Methods

### 2.1 Study Design

The data analyzed in this paper were obtained from persons with HIV enrolled in the Marijuana Associated Planning and Long-term Effects (MAPLE) prospective cohort. Full details of the study have been described elsewhere^34,35^ and also briefly in the following sections. The purpose of the MAPLE study was to investigate the longitudinal effect of marijuana use on neurocognitive functioning and HIV related health outcomes among PWH. Data collection was conducted from 2018 through 2021. Study participants were adults with HIV in Florida recruited at one of three centers in Gainesville (Alachua county), Tampa (Hillsborough county), and Miami (Dade county). All research procedures were approved by the Institutional Review Boards at the University of Florida, Florida International University, and the FDOH. All participants provided informed consent before they participated in the study.

### 2.2 Inclusion and Exclusion Criteria

The MAPLE study recruited both current marijuana users and current non-users. To be eligible for the study as a current marijuana user, study participants had to have confirmed marijuana use by a positive urine THC screen with self-reported marijuana use at least four times per month. Current non-users were defined by a negative urine screen, whose last marijuana use was more than five years ago, and who never used more often than once monthly in their lifetime. The inclusion criteria included being at least 18 years old, living with HIV as confirmed by medical or prescription records, planning to reside in Florida for the next 12 months, English speaking, and being a confirmed current marijuana user or non-user as previously defined. Given the focus of marijuana use quantification, our analysis was restricted to the subsample of current marijuana users.

### 2.3 TLFB Survey Design, Cannabis Use Frequency and Dosage

In this study, a retrospective, calendar-based timeline follow back (TLFB) was used that had been initially developed for alcohol use but modified to measure cannabis consumption. It had been previously found reliable for use with cannabis and other substances, such as cocaine and tabacco^36–38^. Trained MAPLE research assistants used this TLFB to assess patterns of use related to the timing of daily doses, the method of consumption (e.g. joints, blunts, bowls, etc.), the quantity of marijuana flower consumed in grams, and frequency of use over the past 30 days since each study visit.

Participants were first instructed to consider the dose of marijuana they typically used in one sitting or instance of use. For each dose the participant would fill out a dose chart recording the following: the method used (e.g. joint, blunt, pipe), the amount used (e.g. ½ of a joint or 1 whole blunt), the number of hits taken, the number of grams consumed, and the potency of the flower product. Thus, the data was quantified at the individual dosage level for each study participant as opposed to asking participants hypothetically how many joints they could make out of a specified amount of flower or how many hits would they need to consume a joint of a particular size. See figure 1.

In order to help participants accurately estimate the quantity of each method used, they were provided with a picture of different joint sizes and the flower product labeled in grams along with a dollar bill as a reference object during in-person visits. Due to Covid-19, when in-person visits were not possible, telephone visits were conducted and the amounts of marijuana was described in terms of coins: a quarter for 1 gram, a nickel for ¾ of a gram, a penny for ½ of a gram and a dime for ¼ of a gram. See figure 2.

Participants were then asked to think about how their typical doses fit into their typical patterns of use. A pattern might contain multiple doses and must have occurred at least four times in a month to be considered a pattern rather than a deviation. For example, if every weekday a participant took a dose in the morning, then another in the afternoon and two doses in the evening then that would be a weekday pattern. All patterns of use were recorded for participants. See figure 3.

Finally, patterns were embedded into a calendar along with any other methods of use or deviations from typical usage patterns. Anchors dates such as holidays, birthdays or work schedules were used to prompt participant’s recollection of their usual use and any deviations from that use.

Information collected on the TLFB was used to create summary variables such as frequency of use in days per month, quantity used per day, and the number of heavy use days per month, as well as, joint, blunt, bowl and hit size. We examined the distribution of responses, and present the median amounts due to the right skewed distribution of the data.

### 2.4 Statistical Analysis

The primary goal was to calculate the median frequency of use and quantity of grams of flower per day by method of consumption for each participant. Analyses were limited to individuals who had consumed marijuana flower at least once in the past 30 days regardless of whether they were using other cannabis products. We used univariate descriptive statistics and data visualizations to examine sample characteristics and distributions of frequency and quantity of cannabis consumption by method, for example, size of a hit, joint, blunt or bowl to determine the median size consumed by each method, as well as, frequency of use in days per month or quantity consumed in grams per day. Analyses of a particular method of flower consumed included all participants regardless of whether or not they had also used another product types. That is, all analyses of flower consumption included responses provided by participants who exclusively used flower products as well as participants who also used flower along with other products such as concentrates, edibles or vapes. All analyses were done in SAS version 9.4 and R version 4.3.3.

## 3. Results

### 3.1 Demographics, cannabis frequency and quantity used

Table 1 shows the sample socio-demographics. The sample included 253 persons living with HIV who were marijuana flower users. Of the 253 flower users, 208 (82%) of them were exclusively flower users and 45 (18%) used flower along with some combination of other marijuana products such as edibles, vapes, or concentrates. Most participants in the study were adults ≥50 years of age (60.1%), male (54.4%), cis-gender men (52.6%), non-Hispanic Black (66.4%). Nearly all users obtained marijuana from outside of the formal medical system. Among these users, 52% used daily, with a median quantity used of 0.8 grams per day.

### 3.2 Distribution of responses: size of a joint

For participants who reported using joints, the frequency, quantity, size of joints smoked, and number of hits taken during each morning, afternoon and evening time period for the previous 30 days was recorded on their TLFB. Frequency of joint use was 32%. The distribution of their responses for the quantity of marijuana in each joint is given in figure 4. The median size was 0.5 grams with IQR (0.25-1.0).

### 3.3 Distribution of responses: size of a blunt

For participants who reported using blunts, the frequency, quantity, size of blunts smoked, and number of hits taken during each morning, afternoon and evening time period for the previous 30 days was recorded on their TLFB. Frequency of blunt use was 33%. The distribution of the quantity of marijuana in each blunt is given in figure 5. The median size was 1.0 gram with IQR (0.5-1.0).

### 3.4 Distribution of responses: size of a bowl

For participants who reported using bowls, the frequency, quantity, size of bowl smoked, and number of hits taken during each morning, afternoon and evening time period for the previous 30 days was recorded on their TLFB. Frequency of bowl use was 12%. The distribution of the quantity of marijuana in each bowl is given in figure 6. The median size was 0.3 grams with IQR (0.25, 0.5).

### 3.5 Distribution of responses: size of a hit

When recording the consumption of a joint, blunt or bowl, participants were asked for the number of hits taken and the amount of the joint, blunt or bowl consumed. This was done at the individual level for each dose which either formed a pattern of behavior on their TLFB calendar or a deviation. Thus, we were able to estimate the size of a hit for each dose of cannabis. The distribution of the size of hits in grams is given in figure 7. The median size of a hit was 0.063 grams with IQR (0.034-0.125).

### 3.6 Characterization of heavy use

No consensus or published criteria are available to establish what comprises heavy cannabis use days analogous to what is known for that of alcohol. To investigate rates of heavy use, we selected consumption levels of 1 gram, 2 grams, and 3 grams per day in at least one day of the month or every day of the month. The proportion who had at least one heavy use day in a month, or heavy use every day of the month was 30% and 6% when heavy use was defined as 3 grams per day, 43% and 13% for 2 grams per day, and 59% and 23% for 1 gram per day.

## 4. Discussion, limitations and recommendations

In this paper we have presented data from the MAPLE study which was designed to investigate health outcomes associated with cannabis use among PWH. In order to examine the effects of cannabis use upon such health outcomes, we must first develop an accurate method to measure cannabis use. Such a measure is critical for determining the health risks and benefits of cannabis especially when evaluating the outcomes of clinical trials.

This paper describes our method of estimating the daily consumption of cannabis through the use of a multi-dimensional TLFB instrument in order to measure both the frequency in days, and the quantity of flower in grams per day. In terms of frequency, 52% were daily users with a median amount of 0.8 grams per day consumed. We found that the median size of joints and blunts were consistent with what is being reported in the literature. Although, these ranges were derived from studies consisting of a primarily younger sample populations of majority white healthy males, while our population consisted mostly of older, non-Hispanic black males living with a chronic condition and using marijuana primarily for medicinal purposes. However, our estimate for the size of a bowl differed from what is commonly reported in the literature. We found the median size of a bowl to be 0.3 grams, which is smaller than typically reported. The typical size of a hit has been reported as being from 0.01 to 0.1 of a gram, whereas, we found the median size of a hit to be 0.063 grams with IQR (0.091). This is similar to a recent study^39^ in which an the size of a hit was calculated as 0.065 grams with IQR (0.092)^26^ using a unit preference design.

Although daily or near daily cannabis use has been established as one of the best predictors of cannabis use disorder (CUD). On average, adults in the USA with CUD used marijuana 6.2 out of every 10 days over a year and 19% of daily marijuana smokers met the criteria for cannabis dependence^40^. However, there is a scarcity of data concerning just what constitutes heavy cannabis use in terms of quantity. We found almost a quarter of our population used at least 1 gram of cannabis daily and almost a third used 3 grams or more at least one day a month. Further investigation is needed to examine how quantity of cannabis consumed relates to CUD and not just frequency consumed.

### 4.1 Challenges, limitations and recommendations

Conducting the TLFB was a time and resource intensive endeavor for both the patient and the research staff. Although all research staff received detailed training, it is possible that some were better at eliciting more detailed information than others and thus generating higher quality data. Additionally, since only about 10% of the study participants used products other than marijuana flower, we did not attempt to characterize these non-flower products. However, marijuana products and consumption methods other than flower can vary widely and new products are continuing to come to market in states which have legalized marijuana for medical and recreational use. There is a clear need to characterize the frequency of use and quantity consumed of these products. It is worth noting that even in the context of reliable estimation of amount of flower marijuana smoked, substantial variability may exist in the individual’s dose or exposure to physiologically active compounds. This variability can be impacted by a wide range of factors, including smoking behavior (depth and duration of inhalation), lung function and other factors impacting absorption, as well variability in the flower marijuana itself. Future research, particularly when interested in physiological consequences at different doses would benefit from relating self-reported consumption to biomarkers biomarkers for THC and CBD and other metabolites.

### 4.2 Recommendations

Given that there is quite a degree of overlap in the size of a joint and the size of a blunt, a large joint can be considered the size of a small blunt. It could be advantageous to characterize joints and blunts on a spectrum from a small joint (0.25 grams), average joint or small blunt (0.5 grams), large joint or blunt (1.0 gram) and a large blunt (> 1.0 grams). Allowing participants to report use by method on this spectrum would substantially reduce the burden in both time and expense to both the participants and staff. Further, it would allow for more inclusive research of cannabis use, as methods of use may have strong cultural and demographic associations.

## 5. Conclusions

Findings from the present paper extend the existing epidemiological data regarding the prevalence both in terms of frequency of use and quantity of cannabis consumed among older PWH and have implications for the development of future clinical cannabis investigations. The amount of marijuana flower consumed can be difficult to estimate, but our results in PWH who were marijuana flower users are similar to others in defining the average size of a joint, blunt and hit when compared with a young, healthy white population. However, further investigations are needed to establish the optimal methodology needed to estimate the frequency and quantity of cannabis consumed in order to maximize possible accuracy while minimizing the cost of data collection to both research staff and participants in terms of time.

Additionally, over half of this sample consumed greater than one gram of flower per day in the previous month and almost a quarter of the sample population consumed greater than one gram each day of the previous month. Additional research should compare health outcomes and side effects from these different doses and patterns

## Data Availability

All data produced in the present study are available upon reasonable request to the authors

## Availability of data and material

Raw data were generated by the Southern HIV and Alcohol Research Consortium (SHARC) at the University of Florida. Derived data supporting the findings of this study are available from the corresponding author [YW] on request, with approval from the SHARC and appropriate data use agreement.

## Acknowledgments

We would like to thank all the participants and study staff who provided their time to make the Marijuana Associated Planning and Long-term Effects (MAPLE) study possible.

We would also like to thank the following list of organizations for their contributions: Care Resource Community Health Center, Hillsborough County Department of Health, Alachua County Department of Health, and University of Miami Center for AIDS Research (CFAR), and University of Florida Infectious Disease Clinic.

## Author contributions

Conceptualization: RLC, ECP, JV

Funding acquisition: RLC

Project administration YW, GI, GP

Investigation and methodology: RLC, YW, ECP, YL, GP

Data curation: VJ, YL

Formal analysis: YL, DDP

Visualization YL, DDP

Writing – Original draft: DDP

Writing – Review and editing: RLC, SW, YW, ECP, VJ, GI, GP, YL, DDP

All authors approve final version to be published.

